# SCOAT-Net: A Novel Network for Segmenting COVID-19 Lung Opacification from CT Images

**DOI:** 10.1101/2020.09.23.20191726

**Authors:** Shixuan Zhao, Zhidan Li, Yang Chen, Wei Zhao, Xingzhi Xie, Jun Liu, Di Zhao, Yongjie Li

**Affiliations:** MOE Key Lab for Neuroinformation, School of Life Science and Technology, University of Electronic Science and Technology of China, Chengdu, China; West China Biomedical Big Data Center, West China Hospital, Sichuan University, Chengdu, China; Department of Radiology, The Second Xiangya Hospital, Central South University, No.139 Middle Renmin Road, Changsha, Hunan, China; Department of Radiology Quality Control Center, Changsha, Hunan, China; Institute of Computing Technology, Chinese Academy of Sciences, Beijing, China

**Keywords:** COVID-19, convolution neural network, segmentation, lung opacification, attention mechanism

## Abstract

Coronavirus disease 2019 (COVID-19) caused by severe acute respiratory syndrome coronavirus 2 (SARS-CoV-2) has spread worldwide at a rapid rate. As of yet, there is no clinically automated tool to quantify the infection of COVID-19 patients, which is of great significance for judging the disease development and treatment response of patients. Automatic segmentation of lung opacification from computed tomography (CT) images shows excellent potential for this purpose but still faces some challenges, including the complexity and variability features of the opacity regions, the small difference between the infected and healthy tissues, and the noise of CT images. However, due to limited medical resources, it is impractical to obtain a large amount of data in a short time, which further hinders the training of deep learning models. To answer these challenges, we proposed a novel spatial- and channel-wise coarse-to-fine attention network (SCOAT-Net), inspired by the biological vision mechanism, for the segmentation of COVID-19 lung opacification from CT images. SCOAT-Net has a spatial-wise attention module and a channel-wise attention module to attract the self-attention learning of the network, which serves to extract the practical features at the pixel and channel level successfully. Experiments show that our proposed SCOAT-Net achieves better results compared to state-of-the-art image segmentation networks.

## I. Introduction

**T**HE Coronavirus disease 2019 (COVID-19), which is caused by severe acute respiratory syndrome coronavirus 2 (SARS-CoV-2), has become an ongoing pandemic [1]– [4]. As of 9 September 2020, there have been 212 countries with outbreaks, a total of 27,486,960 cases diagnosed, and 894,983 deaths, and the number of infected people continues to increase [5]. Clinically, reverse transcription-polymerase chain reaction (RT-PCR) is the gold standard for diagnosing COVID-19 [6], but it also has the disadvantages of a high false-negative rate [7]–[9] and the inability to provide information about the patient’s condition.

COVID-19 has certain typical visible imaging features, such as lung opacification caused by ground-glass opacities (GGO), consolidation, and pulmonary fibrosis, which can be observed in thoracic computed tomography (CT) images [9]– [11]. Therefore, CT can be used as an essential tool for clinical diagnosis. CT can also directly reflect changes in lung inflammation during the treatment process and is a crucial indicator for evaluating the treatment effect [6]. However, in the course of treatment, the need for repeated inspections leads to a sharp increase in the workload of radiologists. In addition, the assessment of inflammation requires a comparison of the region of lesions before and after treatment. Quantitative diagnosis by radiologists is inefficient and subjective and is difficult to be widely promoted. Artificial intelligence (AI) technology may gradually come to play an important role in CT evaluation of COVID-19 by enabling the evaluation to be carried out more quickly and accurately. AI can also realize the rapid response by integrating multiple functionalities, such as diagnosis [12], segmentation [13], and quantitative analysis [14], assisting doctors in rapid screening, differential diagnosis, disease course tracking, and efficacy evaluation to improve the ability to handle COVID-19. In this study, we focus on the segmentation of COVID-19 lung opacification from CT images.

Benefiting from the rapid development of deep learning [15], many excellent convolution neural networks (CNNs) have been applied to medical image analysis tasks and have achieved the most advanced performance [12], [16], [17]. CNNs can be applied in various image segmentation tasks due to their excellent expression ability and data-driven adaptive feature extraction model. However, the success of any CNN is inseparable from the accurate manual labeling of a large number of training images by medical personnel, so CNNs are not suitable for all tasks. COVID-19 lung opacification segmentation based on CT images is an arduous task that has the following problems. First, in the emergency situation of the COVID-19 outbreak, it is difficult to obtain enough data with accurate labels to train deep learning models in a short time due to limited medical resources. Second, the infection areas in a CT slice show various features such as different sizes, positions, and textures, and there is no distinct boundary, which increases the difficulty of segmentation. Third, due to the complexity of the medical images, the lung opacity area is quite similar to other lung tissues and structures, making it challenging to identify. Several works [18]–[20] have tried to solve these challenges from the perspectives of reducing manual depiction time, using noisy labels, and implementing semi-supervised learning, and have achieved specific results. Our approach in this study is derived from the attention learning mechanism, which makes full use of the inherent extraordinary self-attention ability of CNN to make the network generate attention maps and make the attention vectors in the training process weight the spatial domain feature and channel domain feature. The areas and features activated by the network can diagnose the target area more accurately. Furthermore, a series of studies [21]–[23] have proved the effectiveness of the attention mechanism for classification and segmentation tasks.

The attention mechanism stems from the study of biological vision mechanisms [24], particularly selective attention, a characteristic of human vision. In cognitive neuroscience, it is believed that an individual cannot receive and pay attention to all stimuli due to the bottleneck of information processing. Humans selectively focus on some information while ignoring other visible information. The feature integration theory proposed by Treisman [25] uses a spotlight to describe the spatial selectivity of attention metaphorically. This model points out that visual processing is divided into two stages. In the first stage, visual processing quickly and spontaneously performs low-level feature extraction, including orientation, brightness, and color, from the visual input in various dimensions in a parallel manner. In the second stage, visual processing will locate objects based on the features of the previous stage, generate a map of locations, and dynamically assemble the low-level features of each dimension of the activation area into high-level features. Generally speaking, essential areas attract the attention of the visual system more strongly. Wolfe believes that the attention mechanism uses not only the bottom-up information of the image but also top-down information of the high-level visual organization structure [26], and the high-level information can effectively filter out a large amount of irrelevant information.

This work is inspired by the biological vision mechanism and proposes a novel attention learning method. We use a traditional CNN to complete the extraction of local image features spontaneously. After that, we generate an attention map based on the low-level features of the previous stage to activate the spatial response of the feature, then calculate the attention vector based on the feature interdependence of the activation area to activate the channel response of the feature, and finally complete the reorganization of the high-level features. The attention map and attention vector contain top-down information fed back to the current local features in the form of gating. We call this attention process a coarse-to-fine process, which is a hybrid domain attention mode that includes spatial-wise and channel-wise attention modules.

We believe that the attention learning method proposed above aids the issues faced by segmenting COVID-19 lung opacification. The lung CT slices of patients with pneumonia contain tissue structures easily confused with inflammation areas such as the trachea, blood vessels, emphysema background, and the CNNs method calculations based on the local image, leading inevitably to the overfitting of irrelevant information of the model. We designed the spatial-wise module to generate attention maps in feature extraction, suppressing irrelevant information, and enhancing essential information in the spatial domain. Given the large intra-class differences between opacity regions, the channel-wise module can select and reorganize the spatial domain features.

Based on this method, we propose a spatial and channel-wise coarse-to-fine attention network (SCOAT-Net) and use it to solve the segmentation task of COVID-19 lung opacification. Compared with traditional CNNs, our model recognizes the opacity area better, making it more suitable for complex medical imaging tasks. The contributions of this paper are threefold:

- We propose a novel coarse-to-fine attention network for segmentation of COVID-19 lung opacification from CT images, which utilizes embedded spatial-wise and channel-wise attention modules and achieves state-of-the-art performance (i.e., an average Dice similarity coefficient, or DSC, of 0.8948).
- We use the self-attention method so that the neural network can generate attention maps without external region of interest (ROI) supervision. Furthermore, we use this method to understand the training process of the network by observing the areas that the network focuses on in different stages and increasing the interpretability of the neural network.
- We verify the robustness and compatibility of the SCOAT-Net on different types of CT scans and confirm that it has specific data migration capability. Moreover, it can provide a quantitative assessment of pulmonary involvement, a difficult task for radiologists, and thus enhance the clinical follow-up of patient disease development and treatment response.

## II. Related Works

### A. Segmentation Networks

Deep neural networks (DNNs) have shown excellent performance for many automatic image segmentation tasks. Zhao et al. proposed the pyramid scene parsing network (PSPNet) [27], which introduces global pyramid pooling into the fully convolutional network (FCN) to make the global and local information act on the prediction target together. DeeplabV3 [28], [29] proposed the ASPP (atrous spatial pyramid pooling) module to make the segmentation model perform better on multi-scale objects. U-Net [13] was introduced by Ronneberger et al. based on the encoder-decoder structure that is widely used in medical image segmentation due to its excellent performance. It uses skip connections to connect the high-level low-resolution semantic feature map and the low-level high-resolution structural feature map of the encoder and decoder so that the network output has a better spatial resolution. Oktay et al. [21] proposed the attention gate model and applied it to the U-Net model, which improved the sensitivity and prediction accuracy of the model without increasing the calculation cost. UNet++ [30] uses a series of nested and dense skip paths to connect the encoder and decoder sub-networks based on the U-NET framework, which further reduces the semantic relationship between the encoder and decoder and achieves better performance in liver segmentation tasks.

### B. Artificial Intelligence for COVID-19 based on CT

The segmentation of lung opacification based on CT images is an integral part of COVID-19 image processing, and there are many related works on this topic. Using the lungs and pulmonary opacities manually segmented by experts as standards, Cao et al. [31] and Huang et al. [32] developed a CT image prediction model based on CNNs to monitor COVID-19 disease development, and it showed excellent potential for the quantification of lung involvement. Some studies [33]– [37] trained segmentation models with CT and segmentation templates of abnormal lung cases, which can extract the areas related to lung diseases, making the learning process of pneumonia type classification easier in the next steps. The deep learning model relies on a large amount of data training, and it is impractical to collect a large amount of data with professional labels in a short time. Several research groups [18]–[20] attempted to solve this challenge from the perspectives of reducing manual delineation time, using noisy labels, and implementing semi-supervised learning. VB-Net [18] has a perfect effect on the segmentation of COVID-19 infection regions. The mean percentage of infection (POI) estimation error for automatic segmentation and manual segmentation on the verification set is only 0.3%. In particular, it adopts a human-in-the-loop strategy to reduce the time of manual delineation significantly. Wang et al. [19] proposed noise-robust Dice loss and applied it in COPLE-Net, which surpasses other anti-noise training methods to learn COVID-19 pneumonia lesion segmentation in noisy labels. Inf-Net [20] uses a parallel partial decoder to aggregate high-level features and generate a global map to enhance the boundary area. It also uses a semi-supervised segmentation framework to achieve excellent performance in lung infection area segmentation.

### C. Attention Mechanism

More and more attempts have been focused on the combination of deep learning and visual attention mechanisms, which can be roughly divided into two categories: external-attention mechanisms and self-attention mechanisms. An external-attention mechanism allows the network to learn to generate an attention map during the training process by conducting ROI supervision externally so that the region activated by the network can accurately diagnose disease changes. One study [23], [38] applied this mechanism to the diagnosis of COVID-19 and glaucoma, and the sensitivity was greatly improved. In contrast, a self-attention mechanism does not rely on external ROI supervision but rather exploits the intrinsic self-attention ability of CNN. Self-attention consists of two parts, among which spatial-wise attention [21], [39], [40] redistributes the network’s attention at the pixel level of the feature map to achieve more precise location, and channel-wise attention [41] redistributes the attention at the channel level to instruct the network in selecting practical features. In [42], spatial and channel dimension attention were combined with parallel mode to jointly guide network training, which captured rich contextual dependencies to address the segmentation task. Chen et al. [43] proposed SCA-CNN for the task of image captioning, which incorporated spatial- and channel-wise attention mechanisms. Zhang et al. [22] proposed an attention learning method with the higher layer feature as the attention mask of the lower layer feature, which can achieve the best performance in skin lesion classification.

## III. Method

UNet++ is an excellent image segmentation network which has achieved high-grade performance in medical imaging tasks. It contains dense connections that make the contextual information of different scales closely related. However, although this complicated connection method improves the generalization ability of the model, it also causes information redundancy and weak convergence of the loss function on a small data set. Medical images have the characteristics of high complexity and noise, which cause model overfitting when the amount of training data is insufficient. The SCOAT-Net proposed in this work redesigns the connection structure of UNet++ and introduces the attention learning mechanism. It extracts the spatial and channel features from coarse to fine with only a few added parameters and obtains more accurate segmentation results.

### A. Structure of the Lung Opacification Segmentation Network

Fig. 1 compares the basic structures of UNet++ and the proposed SCOAT-Net. Inheriting the basic structure of UNet++, SCOAT-Net is composed of an encoder and a decoder connected by skip connections. The encoder extracts the information of the semantic level of the image and provides a relatively coarse location, using a max-pooling layer as a down-sampling module. The decoder reconstructs the segmentation template from the semantic information. It uses U-shaped skip connections to receive the corresponding low-level features of the encoder and calculate the final segmentation result. The upsampling module of the decoder uses the bilinear interpolation layer instead of the deconvolution layer to improve the resolution of the feature map. This method dramatically reduces the number of parameters as well as the calculation cost, and it has good performance on small-scale datasets.

**Fig. 1.**
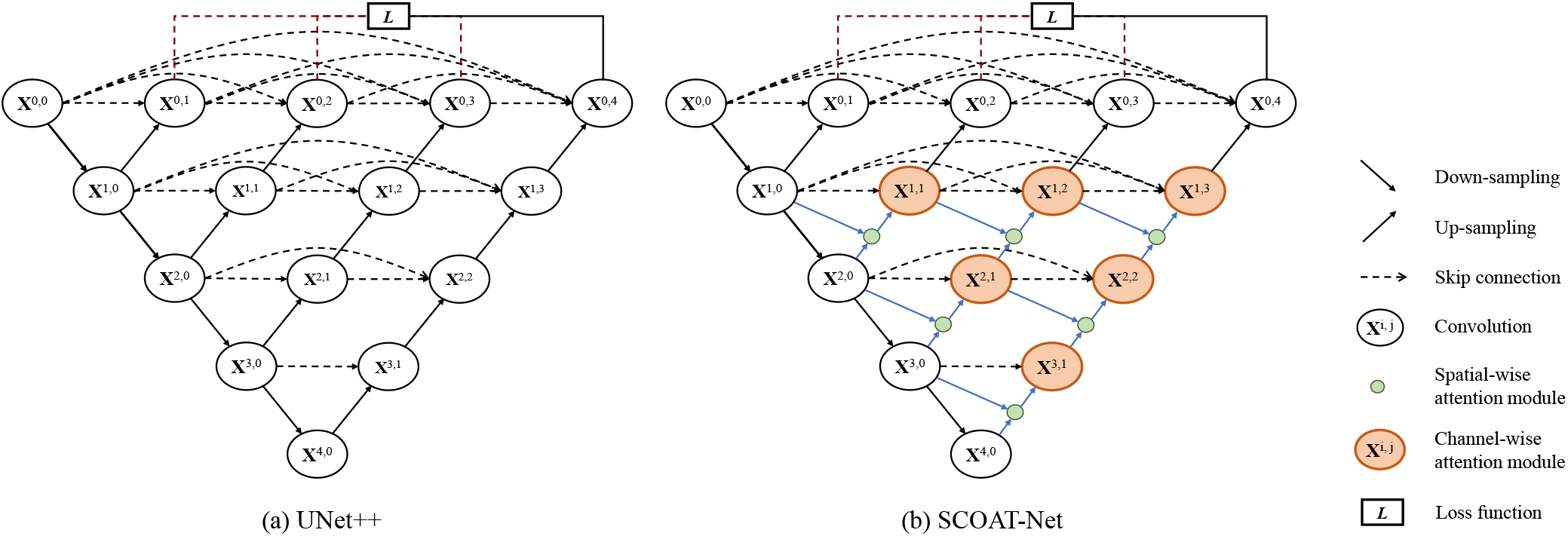
Comparison of UNet++ (a) and the proposed SCOAT-Net (b).

**Fig. 2.**
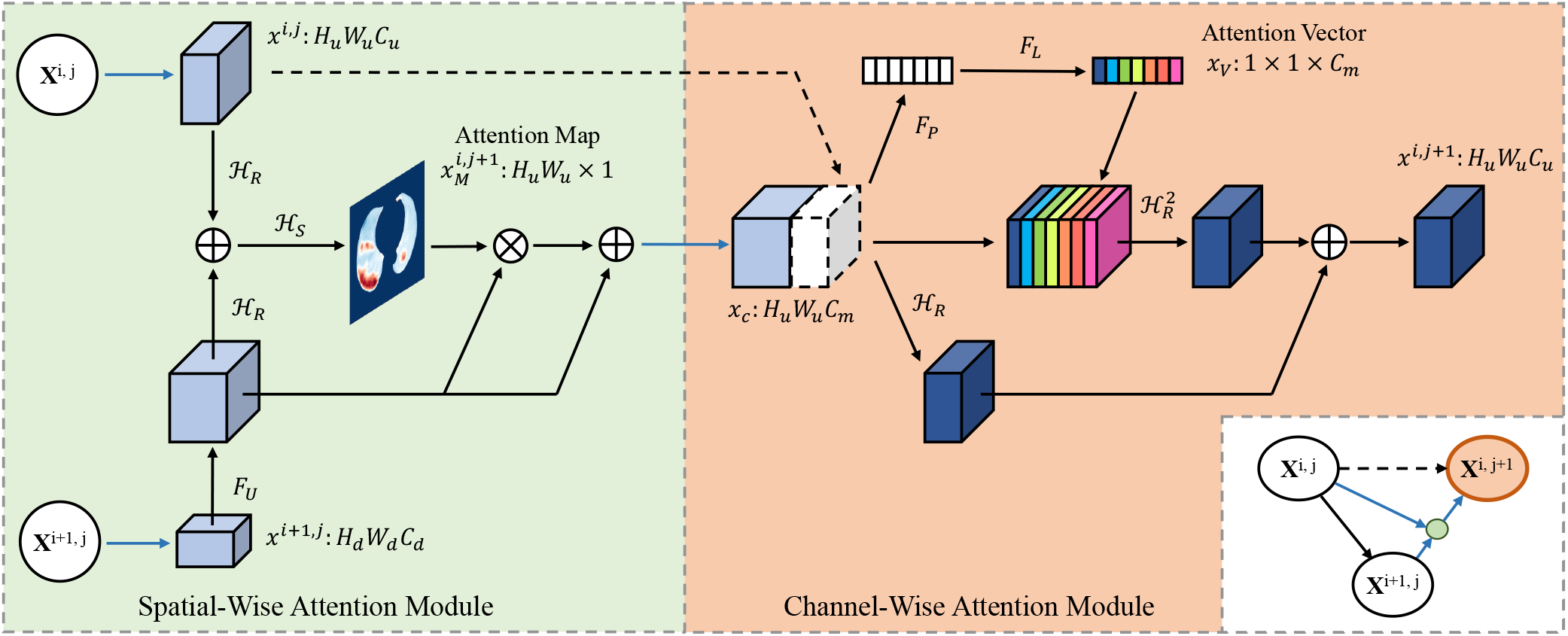
Illustration of spatial-wise attention module and channel-wise attention module.

We reconstruct the connection at the top of the network (except for the bottom layer **X**^0,*j*^) and introduce the attention module. This causes the calculation of the attention mechanism to act on the high-level semantic information and keep the bottom layer of the detailed image information as much as possible, resulting in fine, high-resolution segmentation. The proposed attention module consists of two parts: the spatial-wise attention module and the channel-wise attention module.

We use context feature maps with different resolutions as information of different dimensions for the spatial-wise attention module, as shown in the green circle of Fig. 1, which can combine all the multi-dimensional feature maps extracted by all the filters to calculate the attention map of the image and adjust the target area of the network adaptively. The output of the spatial-wise attention module is contacted with the feature maps of the same layer to enter the channel-wise attention module, as shown in the orange circle. The channel-wise attention module calculates the interdependence between the channels and adaptively recalibrates the information response of the channel. Additionally, in each convolution module, we use the residual block to train our network.

### B. Spatial-Wise Attention

The proposed spatial-wise attention module emphasizes attention at the pixel level, making the network pay attention to the key formation and ignore irrelevant information. Normally, in a CNN, the features extracted by the network change from simple low-level features to complex high-level features with the deepening of the convolutional layers. When calculating the attention map, we can not only use the information of single-layer features but also combine the upper and lower features of different resolutions. The final output of this module is expressed as 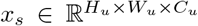, which is given by (1) and (2):

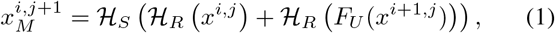

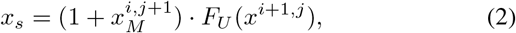

where the function ℋ_*R*_(·) stands for the convolution of size 1 × 1 followed by a batch normalization and a ReLU, used for feature integration. ℋ_*S*_(·) denotes the convolution of size 1 × 1 followed by a batch normalization and a sigmoid activation function, used for feature integration and generation of the attention maps. *F*_*U*_ (·) is the up-sampling operation with a bilinear interpolation function. The input of this module is composed of the upper layer feature 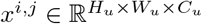 and the lower layer feature 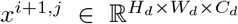, where *x*^*i,j*^ represents the output of each convolution module **X**^*i,j*^. 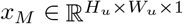 is the attention map generated by this module, which uses the saliency information in the spatial position to weigh the input features to complete the redistribution of the feature attention at the pixel level. The attention map generated by the sigmoid function is normalized between 0 and 1, and the output response will be weakened after point multiplication with the current feature map. Nested structure uses of this method will lead to over-fitting or the degradation of model performance caused by the gradient’s disappearance. To improve this phenomenon, inspired by the ResNet, we add the original features *x*^*i*+1,*j*^ after weighting them by 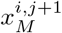, as shown in (2). The final output *x*_*s*_ is sent to the next channel-wise attention module.

### C. Channel-Wise Attention

The input 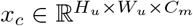 of the proposed channel-wise attention module is obtained by concatenating the spatial-wise attention module’s output *x*_*s*_ with the feature map of the same layer, as in (3):

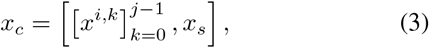

where [·] represents concatenation. 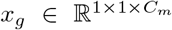 is the channel-wise statistical information calculated by *x*_*c*_ through a global average pooling layer, as in (4), which can reflect the response degree on each feature map.

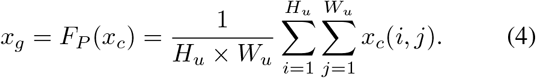

We want the module to adaptively learn the feature channels that require more attention, and we also want it to learn the interdependence between channels. Inspired by the SENet [41], we pass *x*_*g*_ through two fully connected (FC) layers with parameters *ω*_1_ and *ω*_2_ to obtain the attention vector 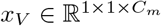 of the channel, as in (5):

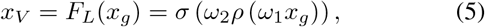

where *ρ*(·) refers to the ReLU activation function, and *σ*(·) refers to the sigmoid activation function. A structure containing two fully connected layers, which reduces the complexity and improves the generalization ability of the model, is adopted here. The fully connected layer of parameter 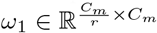 reduces the feature channels’ dimension with reduction ratio *r* (*r* = 16 in this experiment). In contrast, the fully connected layer of parameter 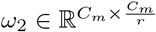 recombines the feature channels to increase its dimension to *C*_*m*_. The attention vector *x*_*V*_ finally weights the input feature map *x*_*c*_, and after the convolution operation completes the feature extraction, it is added to itself to obtain the final output 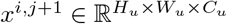, as in (6):

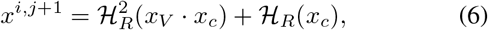

where 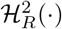 represents the two-layer convolution for feature extraction.

### D. Loss Function

SCOAT-Net has a deep supervision strategy, which can use any one of the segmentation branch outputs (*x*^0,*j*^, *j* ∈ 1, 2, 3, 4) to calculate the loss or use the output of all branches to calculate the average of the loss. The choice depends on the tasks and data. By combining binary cross-entropy (BCE) loss and dice coefficient loss [44], we use a hybrid loss function for segmentation as follows:

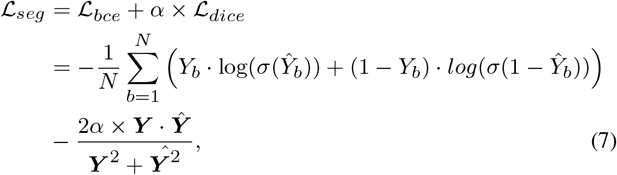

where ***Y*** = {*Y*_1_, *Y*_2_, …, *Y*_*b*_} denotes the ground truths, ***Ŷ*** denotes the predicted probabilities, *N* indicates the batch size, and *σ*(·) corresponds to the sigmoid activation function. This hybrid loss includes pixel-level and batch-level information, which helps the network parameters to be better optimized.

### E. Evaluation Metrics

To evaluate the performance of lung opacification segmentation, we measure the Dice similarity coefficient (DSC), sensitivity (SEN), positive predicted value (PPV), volume accuracy (VA), regional level precision (RLP), and regional level recall (RLR) between the segmentation results and the ground truth, which are defined as follows.

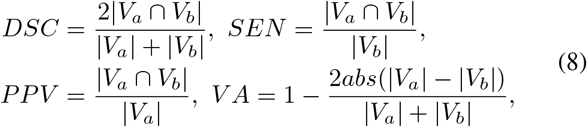

where *V*_*a*_ and *V*_*b*_ refer to the segmented volumes by the model and the ground truth, respectively. In addition to the above voxel-level evaluation indicators, we also design the regional-level evaluation indicators RLP and RLR, as in (9):

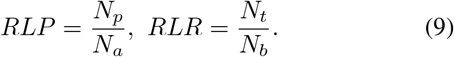

*N*_*a*_ denotes the total number of connected regions of the model prediction result, *N*_*p*_ denotes the number of real opacity regions predicted by the model, denotes the total number of real opacitiy regions, and *N*_*t*_ denotes the number of real opacity regions predicted by the model. If the center of the connected area predicted by the model is in a real opacity region, then we accept that the predicted connected area is correct. We calculate the center of the connected area as:

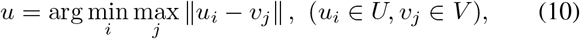

where *U* represents the point set of a single connected area of the prediction result, and *V* represents the point set of its edge.

## IV. Experiment and Results

### A. Data and Implementation

This study and its procedures were approved by the local ethics committees. All methods were performed in accordance with the relevant guidelines and regulations. Written informed consent from the study patients was not required. The data contained 19 lung CT scans of COVID-19 patients obtained using SOMATOM Definition AS and 1117 lung opacification segmentation delineated by radiologists on the single-slice CT. Additionally, we prepared a total of eight lung CT scans of two patients scanned at different times using SOMATOM go.Top, and these scans were used to test the compatibility of our model in different device types. We performed five-fold cross-validation to test the results. The input images were single-layer CT images, which were in the size of 512 × 512 pixels to ensure the high resolution of the result and were normalized before being sent to the network. The sketch templates of the radiologists served as the ground truth, so they were used to calculate the loss function with the final output of the network. We used the gradient descent algorithm with Adam to optimize the loss function that updates the network parameters. The learning rate was set to 0.01, which was multiplied by 0.1 after every ten epoch decays. When the iterative result converged, we adjusted the learning rate to 0.001 for training again. The learning rate decay strategy remained unchanged, and the iteration was set to 50 times. The final results of training in this warm-up [46] method will be slightly improved. All experiments were conducted on an NVIDIA RTX GPU, and the proposed SCOAT-Net was implemented based on a Pytorch framework.

### B. Results on Lung Opacification Segmentation

The aim of this experiment was to evaluate the performance of our proposed SCOAT-Net with different loss functions for lung opacification segmentation. We used six different loss functions, namely MAE [47], IOU [48], BCE, Dice [44], Focal [49], and BCE-Dice, to train the proposed network with the same strategy and hyper-parameters, and the quantitative comparison is listed in Table I. It is evident that IOU, BCE, and Focal had excellent segmentation performance, and their DSCs were the highest. Among them, Focal was superior to IOU and BCE in terms of SEN but slightly inferior in terms of PPV and RLP. It is worth noting that Dice had a more significant performance in terms of SEN and RLR. Dice can predict the entire opacity area better, but it also causes the PPV and RLP performance to decline because it yields more false-positive predictions. The hybrid loss function combining BCE and Dice with parameter *α* (we set *α* = 0.5 in the experiments) produced the best results. Except for SEN and RLR, which were slightly lower than Dice, the other indicators were the best. The box plot shown in Fig. 3 demonstrates the performance of our proposed network with the BCE-Dice loss function. In 19 cases, the model we proposed exhibited excellent performance. The medians of DSC, SEN, and PPV were all higher than 0.9, and the medians of VA, RLP, and PLR were higher than 0.95, even though one or two cases did not achieve excellent results.

**TABLE I.** Quantitative evaluation of SCOAT-Net with defferent loss functions for lung opacification segmentation.

| Loss functions | Results (%) |  |  |  |  |  |
| --- | --- | --- | --- | --- | --- | --- |
|  | DSC | SEN | PPV | VA | RLP | RLR |
| MAE [47] | 85.74 | 84.97 | 83.52 | 91.78 | 82.49 | 84.77 |
| IOU [48] | 88.10 | 86.36 | 90.47 | 94.69 | 92.28 | 88.17 |
| BCE | 88.53 | 86.85 | 91.43 | 94.70 | 92.47 | 89.93 |
| Dice [44] | 86.79 | 89.24 | 85.38 | 90.78 | 86.77 | 92.30 |
| Focal [49] | 87.73 | 87.28 | 87.93 | 93.86 | 88.18 | 88.96 |
| BCE-Dice ($\alpha = 0.5$) | 89.48 | 88.74 | 90.64 | 95.26 | 93.66 | 91.94 |

**Fig. 3.**
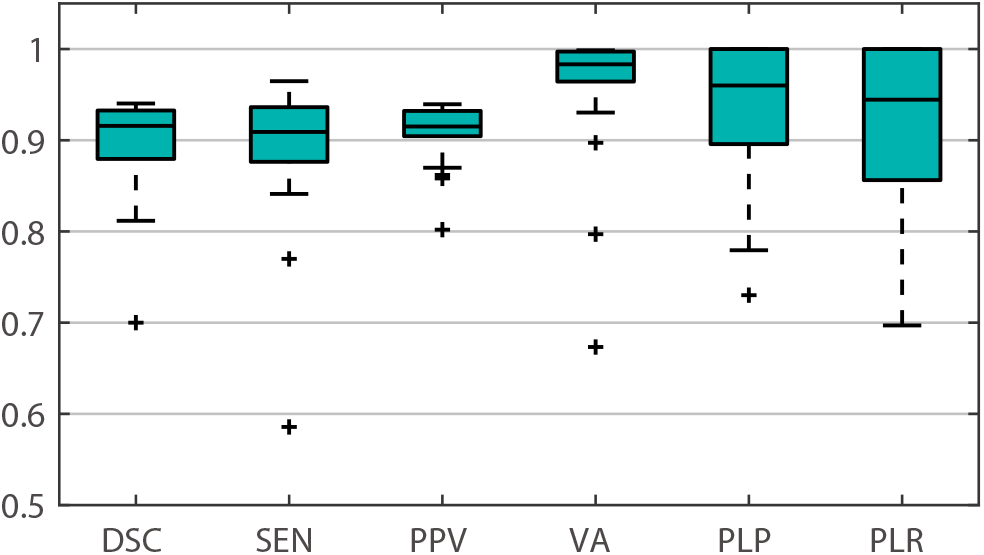
The segmentation performances of SCOAT-Net with BCE-Dice loss function.

### C. Comparison of Different Networks

We compared our proposed SCOAT-Net with other popular segmentation algorithms for lung opacification segmentation. The BCE-Dice loss function was used to train these networks. The quantitative evaluation of these networks was calculated by cross-validation, as shown in Table II. ESPNetv2 had good PPV and RLR, but RLP was extremely low, which shows that the lightweight models could not achieve excellent region-level segmentation results on complex medical image segmentation tasks. DeepLabV3+ achieved an excellent result in Table II, which perhaps results from the good adaptability of its atrous spatial pyramid pooling module designed for semantic segmentation. U-Net, which has an excellent performance in many medical image segmentation tasks, had excellent RLP but the lowest SEN. Although most of the predicted regions were correct, the voxel prediction could not capture all opacity regions. Compared with U-Net, which has a more complex structure and more connections, UNet++ had slightly improved performance in DSC, SEN, and VA, but it had a significant drop in RLP and RLR, which shows that its dense connection improved the model’s generality. However, it did not achieve excellent results on the relatively small dataset used in this work.

**TABLE II.** Quantitative evaluation of defferent networks for lung opacification segmentation. The BCE-Dice loss was used for training.

| Methods | Results (%) |  |  |  |  |  |
| --- | --- | --- | --- | --- | --- | --- |
|  | DSC | SEN | PPV | VA | RLP | RLR |
| PSPNet [27] | 85.08 | 84.95 | 85.84 | 92.32 | 90.71 | 86.06 |
| ESPNetv2 [50] | 86.06 | 83.38 | 89.67 | 91.03 | 75.45 | 90.45 |
| DenseASPP [51] | 85.46 | 83.80 | 89.09 | 91.48 | 87.81 | 88.23 |
| DeepLabV3+ [28] | 87.43 | 86.23 | 89.46 | 94.72 | 91.95 | 90.58 |
| U-Net [13] | 84.72 | 82.23 | 87.90 | 91.13 | 92.00 | 87.68 |
| CE-Net [52] | 78.77 | 78.62 | 81.04 | 80.39 | 79.24 | 82.08 |
| Attention U-Net [21] | 85.64 | 84.87 | 86.69 | 94.94 | 91.44 | 86.30 |
| UNet++ [30] | 84.82 | 83.55 | 86.35 | 94.23 | 89.51 | 85.19 |
| Proposed | 89.48 | 88.74 | 90.64 | 95.26 | 93.66 | 91.94 |

Our proposed SCOAT-Net achieved the best performance among the compared networks. It more effectively identified and segmented the pulmonary opacities by using spatial- and channel-wise attention modules. Fig. 4 shows a visual comparison of the results of each network. In the case #1 to the case #4, SCOAT-Net had the best segmentation performance, not only effectively hitting the target opacity region but also producing the least difference between the segmentation area and the ground truth. However, SCOAT-Net also returned some unsatisfactory segmentation results, as shown in the case #5 of Fig. 4. Most of the models, including our model, failed to predict this tiny opacity region. Although PSPNet makes a valid prediction, it also makes false-positive predictions (e.g., case #1) and false-negative predictions (e.g., case #3), which lead to a decline in the overall performance.

**Fig. 4.**
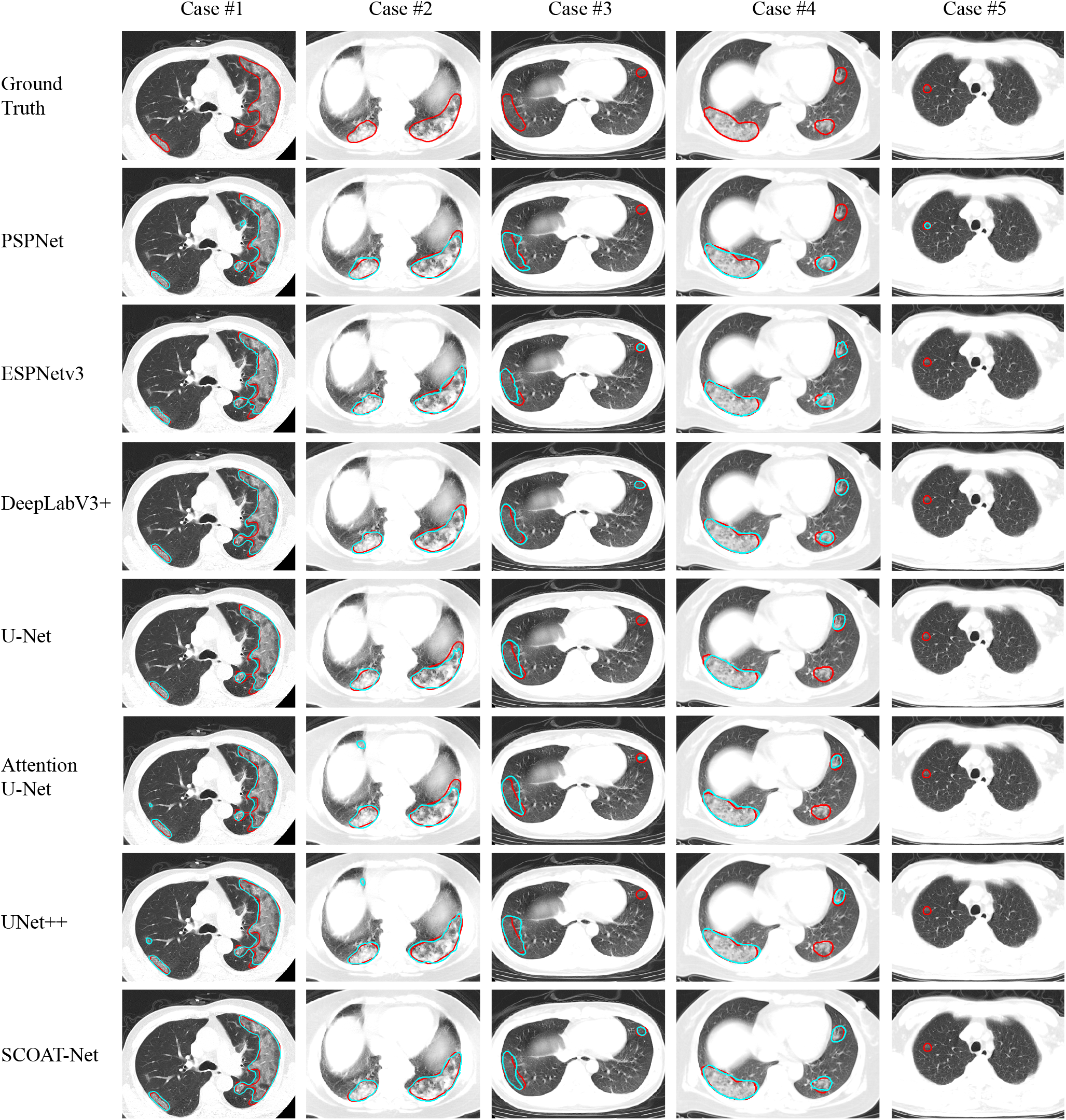
Visual comparison of segmentation performance of different models trained with BCE-Dice loss function. The red curves represent the ground truth, and the cyan curves represent the results of the model.

### D. Effectiveness of the Attention Module

In this experiment, we verified the performance of the attention module on the lung opacification segmentation task. Our SCOAT-Net uses a total of six spatial-wise attention modules, as shown in the green circle in Fig. 1. These modules can adaptively generate attention maps with the focused area information of the network. The early stage of our network is defined as the position that closes to the input and passes fewer convolution layers. The later stage is defined as the position that closes to the output and passes more convolution layers. We selected three different stages of attention maps for display, and the order from the early stage to the late stage is 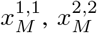, and 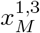, as shown in Fig. 5. For better display, we only show the lung area. We can see that our SCOAT-Net had better performance in lung opacification recognition than UNet++. For example, in the first case, UNet++ identified the interlobular fissure (the yellow arrow area in the lower-left corner) with a specific shape and structure as an opacity region, but SCOAT-Net did not misidentify it. From the attention map of this case, we can see that 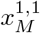 focuses on all the salient areas of the lungs, basically covering all the structures of the lung. Furthermore, 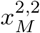 greatly reduces the significant areas, and the attention of the network is more concentrated on restricted regions. By 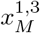, the interlobular fissure area misidentified in the early stage has no longer received the core attention. Additionally, for the opacity region that UNet++ did not recognize (the region indicated by the yellow arrow), SCOAT-Net adequately identified the target area, and on all the attention maps, much attention focused on the target area. As the training phase progressed, the attention regions of SCOAT-Net gradually became smaller. The attention module we designed not only effectively weights the feature map but also further helps us understand the training process of the neural network, which improves its interpretability.

**Fig. 5.**
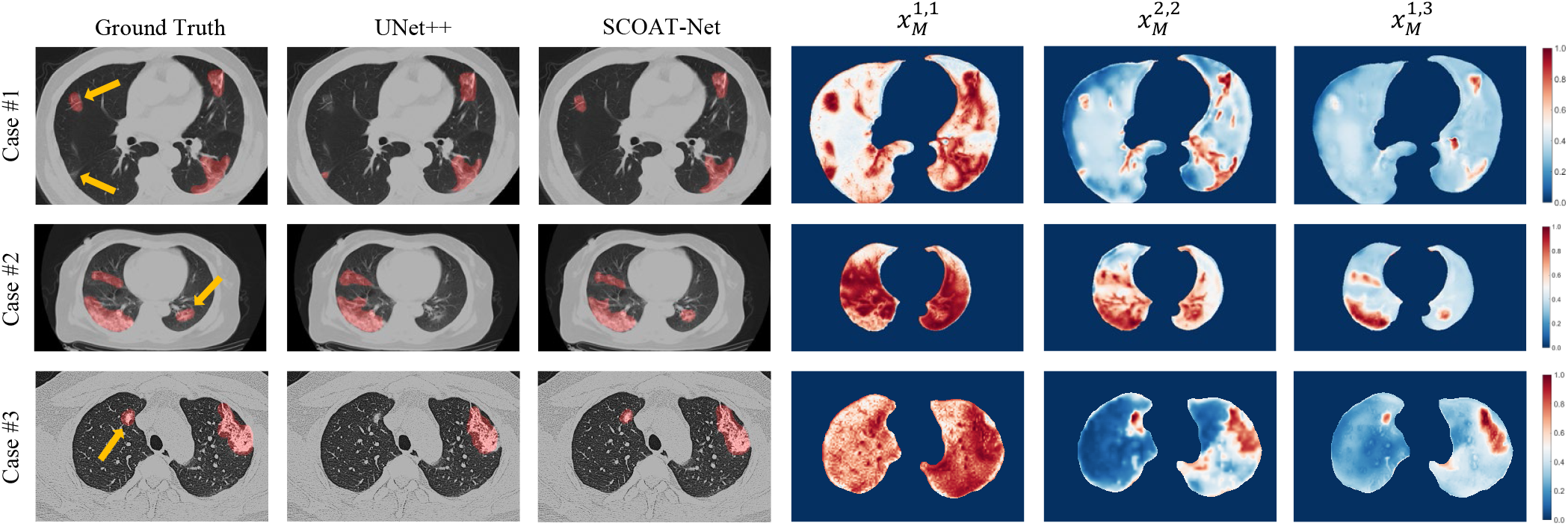
Visualization of the segmentation results and attention maps of our methods on three COVID-19 cases. The red area is the lung opacification segmentation of the ground truth and the models of UNet++ and our SCOAT-Net, and yellow arrows highlight the local differences of the segmentation results.

Furthermore, we also introduced the attention module from other studies into UNet++ and compared the results with that of our SCOAT-Net, as shown in Table III. AttentionV1 uses the attention module of residual attention network [39], AttentionV2 imitates the connection structure of Attention UNet [21], and AttentionV3 uses the pyramid attention module of Wang et al. [40]. Compared with the baseline UNet++, all the networks obtained the significantly improved DSC and RLR. SCOAT-Net and AttentionV2 had outstanding performance in SEN, and SCOAT-Net, AttentionV2, and AttentionV3 had significantly improved RLP. The results show that the attention module can improve the segmentation performance while only increasing a few parameters of the network, especially for the recognition of the target area.

**TABLE III.** Quantitative evaluation of defferent attention module for segmentation. The baseline network is UNet++.

| Methods | Params | Results (%) |  |  |  |  |  |
| --- | --- | --- | --- | --- | --- | --- | --- |
|  |  | DSC | SEN | PPV | VA | RLP | RLR |
| UNet++ | 35.02M | 84.82 | 83.55 | 86.35 | 94.23 | 89.51 | 85.19 |
| AttentionV1 | 53.43M | 86.22 | 85.56 | 87.44 | 93.12 | 90.60 | 87.67 |
| AttentionV2 | 37.63M | 88.01 | 87.14 | 89.28 | 94.88 | 93.79 | 90.96 |
| AttentionV3 | 35.05M | 86.91 | 84.76 | 89.55 | 92.77 | 92.61 | 89.13 |
| Proposed | 38.73M | 89.48 | 88.74 | 90.64 | 95.26 | 93.66 | 91.94 |

### E. Validation on Other Data

We prepared a total of eight lung CT scans of two patients scanned at different times using the CT device SOMATOM go.Top. These scans, which were different from the scans used in training, were used to test the robustness and compatibility of the proposed SCOAT-Net. Fig. 6 presents the lung CT scans of two cases under treatment. COVID-19 is clinically divided into four stages [53]: early stage, progressive stage, peak stage, and absorption stage. The clinical report of the first case shows that it was in the absorption stage at all four time points. From the result of our model, we can see that on both the axial unenhanced and coronal reconstruction CT images, the opacity regions were significantly reduced, which was further verified by the lung opacification volumes (LOVs) displayed on the lower-right corners of the coronal images. The clinical report of the second case shows that the patient was in the early stage at the first time point, the progressive stage at the second time point, and the absorption stage at the third and fourth time points. Our calculated LOV was highest at the second time point, and there was a significant decrease in the third time point, which matched the diagnosis report of the patient. In summary, our proposed SCOAT-Net on cross-modal CT scans was verified, proving that it has better robustness and compatibility. It can provide an objective assessment of pulmonary involvement and therapy response in COVID-19.

**Fig. 6.**
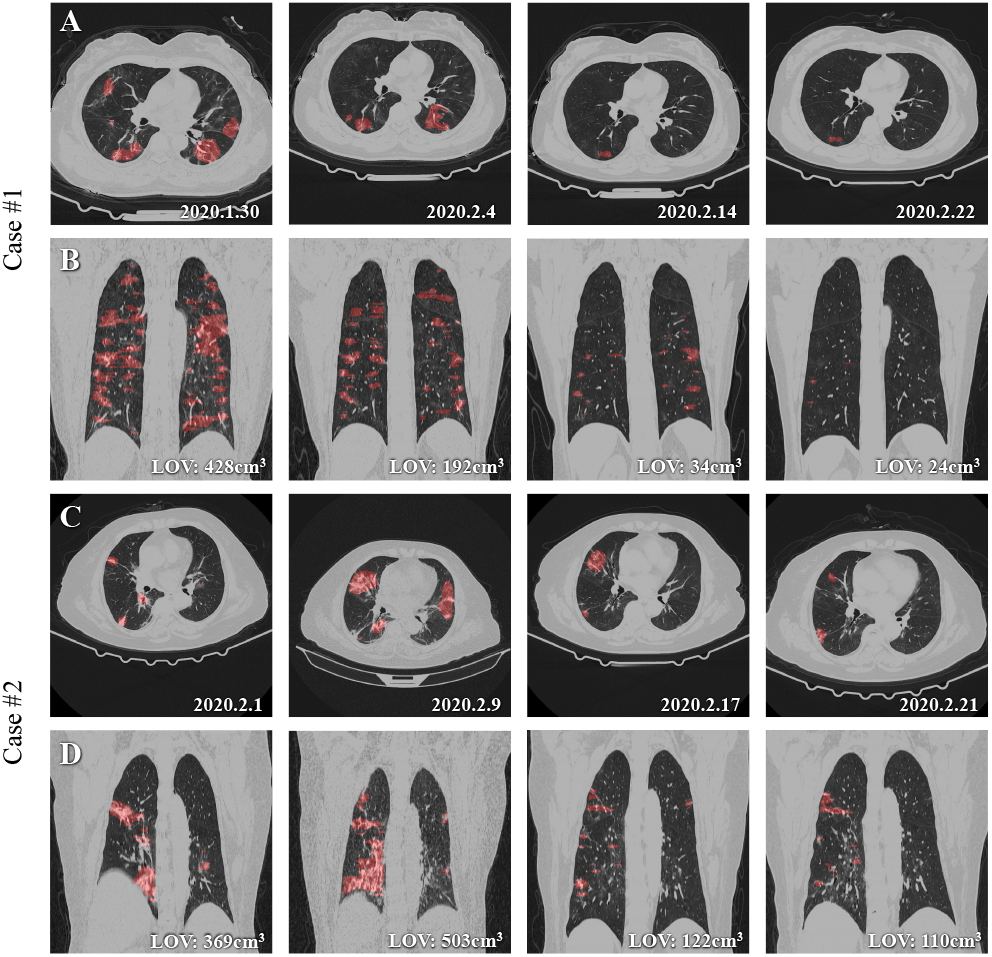
Qualitative evaluation of the results of SCOAT-Net on two cases from other type of CT scan. A and B show the evolution of one COVID-19 case during the 24-day treatment period. C and D show the evolution of another case during the 21-day treatment period. A and C are axial unenhanced chest CT images at four time points (dates are annotated in the lower-right corner of each panel); B and D are the coronal reconstructions at the same time points. The segmentation of pulmonary opacities derived from SCOAT-Net is displayed in red, and the volumetric assessment of our results (i.e., lung opacification volume (LOV)) is annotated in the lower-right corners of the images of B and C.

## V. Discussion and Conclusion

CNNs have been widely used in various medical image segmentation tasks due to their excellent performance [13], [21], [30], [48]. Some networks have been improved from the perspective of connection structure (e.g., U-Net), and others have been improved from the perspective of combining multi-scale features (e.g., PSPNet). These improvements have enhanced the expression ability of the models to a certain extent. However, due to the particularity of medical image-related tasks, only a small amount of data can be obtained, making it impossible to converge when training conventional DNNs, which is a common problem. In addition to augmenting the data [54], some studies show that attention mechanisms can be more effective in enhancing the generalization capacity of models.

The difference between SCOAT-Net and the traditional segmentation network introduces the attention module we designed, which can continuously suppress irrelevant features and enhance useful features in the image space and channel domain during the training process. We applied this network to the task of lung opacification segmentation in COVID-19 cases and achieved better image segmentation performance than state-of-the-art CNNs, as shown in Table 2. It shows that our method has great application potential in complex medical scenarios. Furthermore, we compared the influences of three attention modules in other models and the proposed attention modules in our network on this task. The network incorporating the attention modules has improved performance to varying degrees compared to the baseline network. It is worth mentioning that the attention modules we propose generate a series of attention maps. We can observe the changes of the focused regions at different stages, which contributes to the interpretability of the neural network.

Furthermore, we verified the robustness and compatibility of our network on different types of CT equipment and confirmed that it has excellent data migration capability. Our network can accurately segment lung opacity regions in CT images at different time-points during the treatment. It provides a quantitative assessment of pulmonary involvement, which is a difficult task for radiologists but is essential to the clinical follow-up of patient disease development and treatment response.

However, our network still has shortcomings, as shown in case #5 of Fig. 4. This suggests that we can continue to enhance our network’s recognition of targets of different scales by using multi-scale feature fusion or cascading convolution in different receptive field sizes.

## Data Availability

All data in this study are not available for downloading and re-using.

## Acknowledgment

We thank LetPub for its linguistic assistance during the preparation of this manuscript.

## References

[1] J. T. Wu, K. Leung, and G. M. Leung, “Nowcasting and forecasting the potential domestic and international spread of the 2019-ncov outbreak originating in wuhan, china: a modelling study,” The Lancet, vol. 395, no. 10225, pp. 689–697, 2020.

[2] Z. Wu and J. M. McGoogan, “Characteristics of and important lessons from the coronavirus disease 2019 (covid-19) outbreak in china: Summary of a report of 72 314 cases from the chinese center for disease control and prevention,” JAMA, vol. 323, no. 13, pp. 1239–1242, 04 2020.

[3] H. Shi, X. Han, N. Jiang, Y. Cao, O. Alwalid, J. Gu, Y. Fan, and C. Zheng, “Radiological findings from 81 patients with covid-19 pneumonia in wuhan, china: a descriptive study,” Lancet Infectious Diseases, 2020.

[4] Z. Xu, L. Shi, Y. Wang, J. Zhang, L. Huang, C. Zhang, S. Liu, P. Zhao, H. Liu, L. Zhu et al., “Pathological findings of covid-19 associated with acute respiratory distress syndrome,” The Lancet Respiratory Medicine, 2020.

[5] “Weekly operational update coronavirus disease 2019 (covid-19),” [EB/OL], 2020, https://www.who.int/docs/default-source/coronaviruse/weekly-updates/wou-9-september-2020-cleared-14092020.pdf?sfvrsn=681200132.

[6] Z. Y. Zu, Jiang, P. P. Xu, W. Chen, Q. Q. Ni, G. Lu, and L. J. Zhang, “Coronavirus disease 2019 (covid-19): A perspective from china,” Radiology, pp. 200 490–200 490, 2020.

[7] Y. Fang, H. Zhang, J. Xie, M. Lin, L. Ying, P. Pang, and W. Ji, “Sensitivity of chest ct for covid-19: Comparison to rt-pcr,” Radiology, pp. 200 432–200 432, 2020.

[8] J. F. W. Chan, S. Yuan, K. Kok, K. K. W. To, H. Chu, J. Yang, F. Xing, J. Liu, C. C. Yip, R. W. S. Poon et al., “A familial cluster of pneumonia associated with the 2019 novel coronavirus indicating person-to-person transmission: a study of a family cluster,” The Lancet, vol. 395, no. 10223, pp. 514–523, 2020.

[9] T. Ai, Z. Yang, H. Hou, C. Zhan, C. Chen, W. Lv, Q. Tao, Z. Sun, and L. Xia, “Correlation of chest ct and rt-pcr testing in coronavirus disease 2019 (covid-19) in china: A report of 1014 cases,” Radiology, pp. 200 642–200 642, 2020.

[10] M. Chung, A. Bernheim, X. Mei, N. Zhang, M. Huang, X. Zeng, J. Cui, W. Xu, Y. Yang, Z. A. Fayad et al., “Ct imaging features of 2019 novel coronavirus (2019-ncov),” Radiology, vol. 295, no. 1, pp. 202–207, 2020.

[11] Z. Ye, Y. Zhang, Y. Wang, Z. Huang, and B. Song, “Chest ct manifestations of new coronavirus disease 2019 (covid-19): a pictorial review,” European Radiology, pp. 1–9, 2020.

[12] A. Esteva, B. Kuprel, R. A. Novoa, J. M. Ko, S. M. Swetter, H. M. Blau, and S. Thrun, “Dermatologist-level classification of skin cancer with deep neural networks,” Nature, vol. 542, no. 7639, pp. 115–118, 2017.

[13] O. Ronneberger, P. Fischer, and T. Brox, “U-net: Convolutional networks for biomedical image segmentation,” CoRR, vol. abs/1505.04597, 2015. [Online]. Available: http://arxiv.org/abs/1505.04597

[14] P. Kickingereder, F. Isensee, I. Tursunova, J. Petersen, U. Neuberger, D. Bonekamp, G. Brugnara, M. Schell, T. Kessler, M. Foltyn et al., “Automated quantitative tumour response assessment of mri in neurooncology with artificial neural networks: a multicentre, retrospective study,” Lancet Oncology, vol. 20, no. 5, pp. 728–740, 2019.

[15] Y. Lecun, Y. Bengio, and G. E. Hinton, “Deep learning,” Nature, vol. 521, no. 7553, pp. 436–444, 2015.

[16] D. S. Kermany, M. H. Goldbaum, W. Cai, C. C. S. Valentim, H. Liang, S. L. Baxter, A. Mckeown, G. Yang, X. Wu, F. Yan et al., “Identifying medical diagnoses and treatable diseases by image-based deep learning,” Cell, vol. 172, no. 5, pp. 1122–1131, 2018.

[17] Y. Xie, Y. Xia, J. Zhang, Y. Song, D. Feng, M. J. Fulham, and W. Cai, “Knowledge-based collaborative deep learning for benign-malignant lung nodule classification on chest ct,” IEEE Transactions on Medical Imaging, vol. 38, no. 4, pp. 991–1004, 2019.

[18] F. Shan, Y. Gao, J. Wang, W. Shi, N. Shi, M. Han, Z. Xue, and Y. Shi, “Lung infection quantification of covid-19 in ct images with deep learning.” arXiv: Computer Vision and Pattern Recognition, 2020.

[19] G. Wang, X. Liu, C. Li, Z. Xu, J. Ruan, H. Zhu, T. Meng, K. Li, N. Huang, and S. Zhang, “A noise-robust framework for automatic segmentation of covid-19 pneumonia lesions from ct images,” IEEE Transactions on Medical Imaging, vol. 39, no. 8, pp. 2653–2663, 2020.

[20] D. Fan, T. Zhou, G. Ji, Y. Zhou, G. Chen, H. Fu, J. Shen, and L. Shao, “Inf-net: Automatic covid-19 lung infection segmentation from ct images,” IEEE Transactions on Medical Imaging, vol. 39, no. 8, pp. 2626–2637, 2020.

[21] O. Oktay, J. Schlemper, L. L. Folgoc, M. C. H. Lee, M. P. Heinrich, K. Misawa, K. Mori, S. Mcdonagh, N. Hammerla, B. Kainz et al., “Attention u-net: Learning where to look for the pancreas,” arXiv: Computer Vision and Pattern Recognition, 2018.

[22] J. Zhang, Y. Xie, Y. Xia, and C. Shen, “Attention residual learning for skin lesion classification,” IEEE Transactions on Medical Imaging, vol. 38, no. 9, pp. 2092–2103, 2019.

[23] X. Ouyang, J. Huo, L. Xia, F. Shan, J. Liu, Z. Mo, F. Yan, Z. Ding, Q. Yang, B. Song, F. Shi, H. Yuan, Y. Wei, X. Cao, Y. Gao, D. Wu, Q. Wang, and D. Shen, “Dual-sampling attention network for diagnosis of covid-19 from community acquired pneumonia,” IEEE Transactions on Medical Imaging, vol. 39, no. 8, pp. 2595–2605, 2020.

[24] L. Itti and C. Koch, “A saliency-based search mechanism for overt and covert shifts of visual attention.” Vision Research, vol. 40, no. 10, pp. 1489–1506, 2000.

[25] A. Treisman and G. Gelade, “A feature-integration theory of attention,” Cognitive Psychology, vol. 12, no. 1, pp. 97–136, 1980.

[26] J. M. Wolfe, M. L.-H. Võ, K. K. Evans, and M. R. Greene, “Visual search in scenes involves selective and nonselective pathways,” Trends in cognitive sciences, vol. 15, no. 2, pp. 77–84, 2011.

[27] H. Zhao, J. Shi, X. Qi, X. Wang, and J. Jia, “Pyramid scene parsing network,” arXiv: Computer Vision and Pattern Recognition, 2016.

[28] L. Chen, G. Papandreou, F. Schroff, and H. Adam, “Rethinking atrous convolution for semantic image segmentation,” arXiv: Computer Vision and Pattern Recognition, 2017.

[29] L.-C. Chen, Y. Zhu, G. Papandreou, F. Schroff, and H. Adam, “Encoder-decoder with atrous separable convolution for semantic image segmentation,” in Proceedings of the European conference on computer vision (ECCV), 2018, pp. 801–818.

[30] Z. Zhou, M. M. Rahman Siddiquee, N. Tajbakhsh, and J. Liang, “Unet++: A nested u-net architecture for medical image segmentation,” in Deep Learning in Medical Image Analysis and Multimodal Learning for Clinical Decision Support. Cham: Springer International Publishing, 2018, pp. 3–11.

[31] Y. Cao, Z. Xu, J. Feng, C. Jin, X. Han, H. Wu, and H. Shi, “Longitudinal assessment of covid-19 using a deep learning–based quantitative ct pipeline: Illustration of two cases,” Radiology: Cardiothoracic Imaging, vol. 2, no. 2, p. e200082, 2020.

[32] L. Huang, R. Han, T. Ai, P. Yu, H. Kang, Q. Tao, and L. Xia, “Serial quantitative chest ct assessment of covid-19: Deep-learning approach,” Radiology: Cardiothoracic Imaging, vol. 2, no. 2, p. e200075, 2020.

[33] M. Wang, C. Xia, L. Huang, S. Xu, C. Qin, J. Liu, Y. Cao, P. Yu, T. Zhu, H. Zhu, C. Wu, R. Zhang, X. Chen, J. Wang, G. Du, C. Zhang, S. Wang, K. Chen, Z. Liu, L. Xia, and W. Wang, “Deep learning-based triage and analysis of lesion burden for covid-19: a retrospective study with external validation,” The Lancet Digital Health, vol. 2, no. 10, pp. e506–e515, 2020. [Online]. Available: http://www.sciencedirect.com/science/article/pii/S2589750020301990

[34] O. Gozes, M. Frid-Adar, H. Greenspan, P. D. Browning, H. Zhang, W. Ji, A. Bernheim, and E. Siegel, “Rapid ai development cycle for the coronavirus (covid-19) pandemic: Initial results for automated detection & patient monitoring using deep learning ct image analysis,” arXiv preprint 2003.05037, 2020.

[35] L. Li, L. Qin, Z. Xu, Y. Yin, X. Wang, B. Kong, J. Bai, Y. Lu, Z. Fang, Q. Song et al., “Artificial intelligence distinguishes covid-19 from community acquired pneumonia on chest ct,” Radiology, 2020.

[36] J. Chen, L. Wu, J. Zhang, L. Zhang, D. Gong, Y. Zhao, S. Hu, Y. Wang, X. Hu, B. Zheng et al., “Deep learning-based model for detecting 2019 novel coronavirus pneumonia on high-resolution computed tomography: a prospective study,” MedRxiv, 2020.

[37] S. Jin, B. Wang, H. Xu, C. Luo, L. Wei, W. Zhao, X. Hou, W. Ma, Z. Xu, Z. Zheng et al., “Ai-assisted ct imaging analysis for covid-19 screening: Building and deploying a medical ai system in four weeks,” medRxiv, 2020.

[38] L. Li, M. Xu, X. Wang, L. Jiang, and H. Liu, “Attention based glaucoma detection: A large-scale database and cnn model,” in Proceedings of the IEEE Conference on Computer Vision and Pattern Recognition, 2019, pp. 10 571–10 580.

[39] F. Wang, M. Jiang, C. Qian, S. Yang, C. Li, H. Zhang, X. Wang, and X. Tang, “Residual attention network for image classification,” in Proceedings of the IEEE conference on computer vision and pattern recognition, 2017, pp. 3156–3164.

[40] W. Wang, S. Zhao, J. Shen, S. C. Hoi, and A. Borji, “Salient object detection with pyramid attention and salient edges,” in Proceedings of the IEEE Conference on Computer Vision and Pattern Recognition, 2019, pp. 1448–1457.

[41] J. Hu, L. Shen, S. Albanie, G. Sun, and E. Wu, “Squeeze-and-excitation networks,” IEEE Transactions on Pattern Analysis and Machine Intelligence, pp. 1–1, 2019.

[42] J. Fu, J. Liu, H. Tian, Y. Li, Y. Bao, Z. Fang, and H. Lu, “Dual attention network for scene segmentation,” in Proceedings of the IEEE Conference on Computer Vision and Pattern Recognition, 2019, pp. 3146–3154.

[43] L. Chen, H. Zhang, J. Xiao, L. Nie, J. Shao, W. Liu, and T. Chua, “Sca-cnn: Spatial and channel-wise attention in convolutional networks for image captioning,” pp. 6298–6306, 2017.

[44] F. Milletari, N. Navab, and S.-A. Ahmadi, “V-net: Fully convolutional neural networks for volumetric medical image segmentation,” in 2016 fourth international conference on 3D vision (3DV). IEEE, 2016, pp. 565–571.

[45] D. P. Kingma and J. Ba, “Adam: A method for stochastic optimization,” arXiv: Learning, 2014.

[46] A. Gotmare, N. S. Keskar, C. Xiong, and R. Socher, “A closer look at deep learning heuristics: Learning rate restarts, warmup and distillation,” arXiv: Learning, 2018.

[47] C. J. Willmott and K. Matsuura, “Advantages of the mean absolute error (mae) over the root mean square error (rmse) in assessing average model performance,” Climate research, vol. 30, no. 1, pp. 79–82, 2005.

[48] H. Huang, L. Lin, R. Tong, H. Hu, Q. Zhang, Y. Iwamoto, X. Han, Y.-W. Chen, and J. Wu, “Unet 3+: A full-scale connected unet for medical image segmentation,” in ICASSP 2020-2020 IEEE International Conference on Acoustics, Speech and Signal Processing (ICASSP). IEEE, 2020, pp. 1055–1059.

[49] T.-Y. Lin, P. Goyal, R. Girshick, K. He, and P. Dollár, “Focal loss for dense object detection,” in Proceedings of the IEEE international conference on computer vision, 2017, pp. 2980–2988.

[50] S. Mehta, M. Rastegari, L. Shapiro, and H. Hajishirzi, “Espnetv2: A light-weight, power efficient, and general purpose convolutional neural network,” in Proceedings of the IEEE conference on computer vision and pattern recognition, 2019, pp. 9190–9200.

[51] M. Yang, K. Yu, C. Zhang, Z. Li, and K. Yang, “Denseaspp for semantic segmentation in street scenes,” in Proceedings of the IEEE Conference on Computer Vision and Pattern Recognition, 2018, pp. 3684–3692.

[52] Z. Gu, J. Cheng, H. Fu, K. Zhou, H. Hao, Y. Zhao, T. Zhang, S. Gao, and J. Liu, “Ce-net: Context encoder network for 2d medical image segmentation,” IEEE transactions on medical imaging, vol. 38, no. 10, pp. 2281–2292, 2019.

[53] H. Li, S. Liu, H. Xu, and J. Cheng, “Guideline for medical imaging in auxiliary diagnosis of coronavirus disease 2019,” Chin J Med Imaging Technol, vol. 36, no. 3, pp. 321–331, 2020.

[54] A. Zhao, G. Balakrishnan, F. Durand, J. V. Guttag, and A. V. Dalca, “Data augmentation using learned transformations for one-shot medical image segmentation,” arXiv: Computer Vision and Pattern Recognition, 2019.

